# Poverty trajectories and child and mother wellbeing outcomes in Ireland: findings from an Irish prospective cohort

**DOI:** 10.1101/2023.12.07.23299656

**Authors:** David J O Driscoll, Elizabeth Kiely, Linda M O Keeffe, Ali S Khashan

**Author notes:** **David J O Driscoll** Roles: Conceptualization, formal analysis, investigation, methodology, project administration, validation, writing – original draft preparation. **Elizabeth Kiely** Roles: Conceptualization, methodology, investigation, project administration, supervision, visualization, writing – review & editing. **Linda O Keeffe** Roles: Conceptualization, methodology, formal analysis, investigation, project administration, supervision, visualization, writing – review & editing. **Ali Khashan** Roles: Conceptualization, methodology, formal analysis, investigation, project administration, supervision, visualization, writing – review & editing.

## Abstract

**Background:** Poverty is associated with poor outcomes, yet exposure to distinct poverty trajectories in early childhood is not well understood.

**Objective:** To understand the prevalence of different trajectories of household poverty and their association with mid-childhood and primary-parent(mother) indicators of physical health and psychopathology in Ireland.

**Methods:** We used a nationally representative, prospective cohort (Growing-up-in-Ireland– Infant-Cohort). Household poverty included lowest third income decile, subjective poverty and material deprivation when children were aged 9-months(m),3,5,9-years(y). We used group based multi-trajectory-cluster-modelling to classify trajectories of poverty. Using multivariate logistic regression, adjusted with separate child and mother confounders, we assessed the association of poverty trajectories from 9m to 9y with child outcomes (overweight, any longstanding illness and psychopathology) at age 9y and the same poverty trajectories over the same 9y period with mother outcomes (overweight, any longstanding illness and depression).

**Results:** Of 11,134 participants, four trajectories were identified: never-in-poverty (43.1%), material/subjective>monetary-poverty (16.1%), monetary>material-poverty (25.6%), and persistent-poverty (15.2%). Children in persistent-poverty compared with never in poverty experienced higher odds of being overweight at 9y (adjusted odds ratio aOR 1.70,95% CI 1.34, 2.16), having a longstanding illness (aOR 1.51,95% CI 1.20, 1.91) and psychopathology (aOR 2.06,95% CI 1.42, 2.99). The outcomes for primary parent (99.7% were mothers) had higher odds of being overweight (aOR 1.49,95% CI 1.16, 1.92), having a longstanding illness (aOR 2.13,95% CI 1.63, 2.79), and depression (aOR 3.54,95% CI 2.54, 4.94).

**Conclusions:** Any poverty trajectory was associated with poorer psychopathology and physical wellbeing in late childhood for children and their mothers in Ireland.

**Take home message:** *What is already known on this subject?:* Poverty is understood as having both objective (e.g. low household income) and subjective dimensions (e.g., perception of being poor relative to others, poverty related anxiety). Evidence highlights the association between child poverty exposure and child and mother wellbeing outcomes.

*What this study adds?:* Our study examined predicted trajectories of multi-dimensional poverty and child and mother outcomes using a large national cohort. We identified that any poverty trajectory in childhood had a higher odds of being associated with poor well-being outcomes.

*How this study might affect research, practice or policy?:* This study highlights the need for strategies to ameliorate any poverty exposure as all poverty trajectories resulted in poorer wellbeing outcomes. Strategies should be considered to ameliorate any poverty exposure in early childhood (e.g., school based resourcing).

## Introduction

Adverse childhood experiences (ACEs) are a public health concern (Bellis et al., 2019; Hughes et al., 2017). Early life experience of ACE’s (e.g., violence, abuse, neglect) are associated with poorer child outcomes (e.g., education attainment, mental and physical ill-health) (Crouch et al., 2019). Poverty is an ACE and there is growing evidence that children in poverty also experience other clustered childhood adversities (Adjei et al., 2022; Lee et al., 2021). Moreover, children in poverty may experience adverse outcomes in childhood and during the life course into adulthood (e.g., chronic mental and physical illness, early mortality) (Lee et al., 2021).

Poverty is often assessed using monetary metrics (e.g., <60% median income, lowest decile) (Fitzsimons et al., 2017; Green et al., 2018; Lai et al., 2019; Lee and Zhang, 2022). Yet, poverty is challenging to define, conceptualise and measure, especially in a high-income country (Cooper et al., 2012; Laderchi et al., 2003; Streeten, 1998). Hence there is a need for a broader definition encompassing a multi-dimensional approach to poverty in public health research and health outcomes (The Lancet Child Adolescent Health, 2019). Poverty may be subjectively experienced (e.g., inability to pay bills or anxiety about them) or it may be a measure of inability to afford items that in turn contribute to material deprivation (e.g., inability to heat a home, to buy food, to buy two pairs of shoes etc.). It is unclear from a trajectory perspective if certain children are experiencing different types of poverty trajectories (e.g., subjective, material, monetary, or persistent poverty) and whether there are differences in outcomes in later childhood. Group-based multi-trajectory modelling has been used to identify such trajectories (e.g., household poverty, childhood adversity) yet there is a need to use a broadened definition of child poverty in the literature (Adjei et al., 2022; Lai et al., 2019). By isolating clusters of children who share common trajectories of poverty across childhood, we may be able to target them more effectively for developmentally appropriate public policy interventions (Lee et al., 2023; Wickham et al., 2016).

Addressing socio-economic inequality is challenging. A public policy initiative to ameliorate the worst impacts of educational inequality for children in families from lower socioeconomic backgrounds has been in place in Ireland since 2005 called the Delivering Equality of Opportunity in Schools (DEIS) programme (“DEIS Delivering Equality of Opportunity In Schools,” 2020). It provides additional supports to schools (primary school (age 5-12 years) and secondary school (age 13-18 years) that have a higher prevalence of children from lower socioeconomic backgrounds. This support is in the form of grant aids, funding, staffing, home schooling community liaison services, a school meal programme, priority access to professional development supports and a school completion programme designed to discourage early school leaving. There is a need to understand if a poverty trajectory outcome has different stratification by DEIS school attendance for both a child and/or their parents. Prior literature focuses on DEIS school efficacy with regard to academic potential (“DEIS Delivering Equality of Opportunity In Schools,” 2020; Fenwick et al., 2022; Fleming and Harford, 2023). An educational system through the state has a broader responsibility to offset the worst disadvantaged families to improve child and parent outcomes (e.g., behaviour, emotions, longstanding illness, obesity).

We hypothesise that children in a high income country (e.g., Ireland) experience distinct trajectory clusters of household poverty during the first 9 years of childhood and that these trajectories are associated with child and mother health outcomes. Moreover, that attendance at a school with additional resources (e.g., DEIS) for students experiencing household poverty may change this association.

Using a nationally representative prospective cohort, we identified trajectories of multidimensional household poverty during the childhood life course. We then examined the association between these trajectories from 9 months to 9 years of age and (i) child outcomes at 9 years of age (psychopathology, overweight and longstanding illness), (ii) parent (mother) outcomes (mean (SD) age was 41.1 (5.3) years) after 9 years when their child is 9 years of age (depression, overweight, and longstanding illness), and (iii) child sex and attendance at a resourced school (e.g., DEIS) for families experiencing poverty stratification analysis.

## Methods

### Data

Data was used from the Infant Cohort as part of the Growing up in Ireland (GUI) study (Quail et al., 2011a, 2011b). The Infant Cohort is estimated to represent 1-in-9 children born in 2008 in Ireland (n= 11, 194). Using the national child benefit register as a sampling frame, the Infant Cohort was recruited in 2008 (at 9 months (n= 11 134)) and followed up at 3 (n=9 793), 5 (n=9 001), and 9 (n=8 032) years of age. Data at 7 years of age was excluded in this study as it was a postal questionnaire instead of an in-person interview and it did not contain the relevant exposure variables. The primary care giver (PCG) (referred to as mother now as the cohort at 9 months and 9 years was primarily mother >99.7%) were interviewed face-to-face using computer-aided personal interviewing by trained interviewers. Quail et al, provide a detailed description of the study procedure (Quail et al., 2011a, 2011b). The teacher of each child participant was interviewed at 5 years and 9 years of age in the Infant Cohort.

### Household poverty exposure trajectories

Using a similar approach to a GUI report, the inputs to compute predicted household poverty exposure trajectories included three components (monetary poverty, subjective sense of poverty and material poverty) from each wave of the Infant Cohort (age 9 months, 3, 5, 9 years) (**Figure S1, Table S1, Table S2**) (Maître et al., 2021).

### Child Outcomes at 9 years of age

#### Strengths and Difficulties Questionnaire (SDQ)

The SDQ was devised to identify current and early signs of emotional difficulties, hyperactivity, conduct behaviour, peer problems and prosocial issues in children (Goodman, 1997). Each question item has a three-point scale, ‘*not true*’, ‘*somewhat true*’, or ‘*certainly true*’ and each subscale has a total score (0-10). The SDQ has been validated in many jurisdictions (Heiervang et al., 2007; Muris et al., 2003). The SDQ has strong psychometric properties and is used in many longitudinal cohorts (Goodman, 2001; Stone et al., 2010; Van Roy et al., 2008). We were interested in total difficulties scores. We categorised the continuous scale into a binary variable (≥ 17). The mother and teacher completed the SDQ at 9 years of age.

#### Body Mass Index (BMI)

An interviewer measured the weight and height of each child at 9 years of age to derive body mass index. A Leicester portable height measure was used to obtain height (to nearest millimetre). A SECA 761 flat mechanical scales (class IIII medically approved scales) was used to obtain weight (to nearest kg). BMI=weight(kg)/height^2^(m) was categorised based on international guidelines as per World Health Organisation (WHO Consultation on Obesity (1999: Geneva and Organization, 2000) (normal <25, overweight 25-30, and obesity>30). A score of overweight or obese was categorised as overweight for poor health outcome.

#### Longstanding illness

At 9 years of age, a mother was asked if their child had, “any on-going chronic physical or mental health problem, illness or disability?”. A “yes” response indicated a poor health outcome.

### Mother Outcomes at after 9 years when child was 9 years of age (mother mean (SD) age was 41.1 (5.3) years)

#### Centre for epidemiological studies depression scale (8-item) (CESD-8)

The CESD-8 is a shorted version of the widely used self-report (CES-D) screen tool for depression (Burns et al., 2018; Dang et al., 2020). It has been shown to discriminate children and adults with depressive disorder. The short (8 item) scale (CESD-8) is highly correlated with the full 20-item (r=0.93). Participants are asked 8 questions and a composite score ≥7 indicates a depressive score (see **Table S3** for CESD-8 questions).

#### Body Mass Index (BMI)

Similar to child BMI outcome method, a mother’s BMI was measured.

#### Longstanding illness

A mother was asked about themselves if they had, “any on-going chronic physical or mental health problem, illness or disability?”. A “yes” response indicated a poor health outcome.

### Potential confounders

Each potential confounder was measured prior to poverty trajectory exposure. These are illustrated in two directed acyclic graphs (child outcomes **Figure S3**, and mother outcomes **Figure S4**). When assessing child outcomes, we considered child sex, mother age, mother education (degree or higher qualification or no degree), mother health status at 9 months (chronic illness, BMI), mother depression status at 9 months (raised depression score), homeowner, household social class and composition, and child birthweight (<2500g (i.e., low birth weight) or >2500g) and pregnancy gestation (<37 weeks (i.e., preterm) or >37 weeks) (**Figure S3)**. When assessing mother outcomes, we considered child sex, mother age, mother education (degree or higher qualification or no degree), mother health status at 9 months (chronic illness, BMI), mother depression status at 9 months (raised depression score), home-owner, household social class and composition (**Figure S4)**.

### Statistical analysis

#### Group based trajectory

The optimal number of trajectory group memberships was determined by estimating models with increasing latent class trajectories (group based modelling of longitudinal data) (van de Schoot et al., 2017). The Bayesian Information Criterion (BIC) approximation to the Bayes factor between two adjacent models (e.g., 2 and 3 groups) was chosen based on lowest BIC value pending adequacy of the model (van de Schoot et al., 2017). Cubic polynomials were used to represent trajectories. If the higher order term was not statistically significant (p<0.05), lower order terms were refitted to the model (Nagin, 2005). The adequacy of the model was determined by average posterior probabilities (APPs) greater than 70% across trajectory groups indicating adequate classification (Nagin, 2005). The degree of classification accuracy of placing participants into a trajectory based on their posterior probabilities was determined by relative entropy (>0.8 indicates better classification certainty) (Herle et al., 2020; Nylund-Gibson et al., 2023). Finally, mismatch is the difference between the estimated probability of group membership and the proportion of individuals classified in that group based on the highest posterior probability. The correspondence between these two probabilities decreases as assignment error increases (Nagin, 2005). Therefore, a difference close to zero suggests an adequate fit (Lennon et al., 2018; Nagin, 2005). We tabulated model selection results and model adequacy for transparency. This analysis was performed using Stata TRAJ package. Full-information maximum likelihood (FIML) was used to account for missing data and longitudinal weights were used to account for representative bias and attrition.

#### Primary analyses

We described the characteristics of the cohort participants across poverty trajectories. The association between poverty trajectories and child outcomes were examined using multivariable logistic regression and presented as odds ratio (ORs) with 95% confidence intervals (CIs). Unadjusted and adjusted models were reported. Similarly, unadjusted and adjusted models of the association between poverty trajectories and mother outcomes were reported.

#### Sensitivity analyses

We described in tabulation form input exposure variables and group based trajectory subgroups to illustrate construct validity. We repeated our main analysis and examined if the effect of a poverty trajectory differs by DEIS school attendance (i.e., extra resourced school (DEIS) in an area of deprivation) in child and mother outcomes. We again repeated our main analysis and examined if the effect of poverty trajectory differs by child sex in child and mother outcomes. To assess robustness of results, we repeated the main analysis using multiple imputation chained equation (25 imputed data sets with results pooled using Rubin’s rules), and imputed the missing child and mother outcomes. As the outcomes investigated are common health outcomes, we ran a sensitivity analysis by reporting the incidence risk ratio for child and mother outcomes using Poisson regression. All data analysis was carried out using the statistical software package Stata (v.18).

## Results

### Poverty trajectories and demographics of Infant Cohort

A total of 11,134 participants were included in the group-based trajectory analysis provided that input poverty exposures were present in at least one wave. **Table S4** illustrates that a fourgroup trajectory was the best fit with reasonable entropy (i.e., >0.8). A higher order subgroup trajectory did not provide qualitatively distinct groups. The four group trajectory had adequate membership, an APP>0.70 and odds of correct classification (OCC) was much higher than five. The persistent poverty group had an OCC>57.12 (**Table S5**). **Table S6** illustrates that each group trajectory breakdown of input exposures and provide qualitative validity of group membership. Four poverty trajectory clusters were identified: never in poverty (n=4 804, 43.1%), vulnerable to poverty group 1 (n=1 791, 16.1%), vulnerable to poverty group 2 (n=2 851, 25.6%) and persistent poverty (n=1 688, 15.2%) (**Table 1**). Vulnerable to poverty group 1 had moderate subjective/material poverty and low monetary poverty (**Figure S3)**. Vulnerable to poverty group 2 had both moderate perceived/material and monetary poverty (**Figure S2**). There was relatively equal male to female children, and most primary care givers at 9 months were a parent and female indicating mother (**Table 1**). The persistent poverty group compared to never in poverty group had fewer mothers with a degree (n=224, 13.3%), lower home ownership (n=578, 34.2%), more one parent households with 2+ children (n=342, 20.3%) and higher never worked at all family social class (n=619, 36.7%). The persistent poverty group compared to never in poverty group for child outcomes had higher total difficulties as reported by teacher (n=246, 14.6%) and mother (n=147, 8.7%), higher levels of being overweight (n=320, 19%) and having a longstanding illness (n=303, 18%). The persistent poverty group compared to never in poverty group for mother outcomes had higher depression scores (n=215, 12.7%), higher levels of being overweight (n=608, 36%) and having a longstanding illness (n=326, 19.3%).

**Table 1:** Study description of poverty dynamic exposures and outcomes in the Irish Growing Up in Ireland Cohort Study up to age 9 years (Wave 5)

|  | **Never in Poverty**<br>(n=4,804)<br>n (%) | **Vulnerable to Poverty 1**<br>(n=1,791)<br>n (%) | **Vulnerable to Poverty 2**<br>(n=2,851)<br>n (%) | **Persistent Poverty**<br>(n=1,688)<br>n (%) |
| --- | --- | --- | --- | --- |
| **Child co-variates** |  |  |  |  |
| Gender (female) | 2,324 (48.4) | 893 (49.9) | 1,422 (49.9) | 816 (48.3) |
| Relationship to PCG (parent) | 4,804 (100) | 1,790 (99.9) | 2,851 (100) | 1,687 (99.9) |
| Low Birth Weight <2500g | 253 (5.3) | 98 (5.5) | 187 (6.6) | 134 (7.9) |
| Preterm<37weeks | 287 (6) | 106 (5.9) | 200 (7) | 144 (8.5) |
| Extra resourced school (DEIS) in an area of deprivation | 328 (6.8) | 200 (11.2) | 424 (14.9) | 295 (17.5) |
| **PCG co-variates at 9m** |  |  |  |  |
| Gender (female) | 4,784 (99.6) | 1,786 (99.7) | 2,846 (99.8) | 1,680 (99.5) |
| *PCG age categories* |  |  |  |  |
| Less than 26 years | 359 (7.5) | 226 (12.6) | 906 (31.8) | 509 (30.2) |
| 27-30 years | 918 (19.1) | 352 (19.7) | 631 (22.1) | 372 (22) |
| 31-35 years | 2,063 (42.9) | 711 (39.7) | 748 (26.2) | 431 (25.5) |
| 36-39 years | 1,143 (23.8) | 360 (20.1) | 412 (14.5) | 263 (15.6) |
| 40+ | 321 (6.7) | 142 (7.9) | 154 (5.4) | 113 (6.7) |
| Degree - Yes | 2,507 (52.2) | 745 (41.6) | 555 (19.5) | 224 (13.3) |
| Chronic health problem - Yes | 415 (8.6) | 260 (14.5) | 289 (10.1) | 306 (18.1) |
| Raised depression score - Yes | 30 (0.6) | 38 (2.1) | 42 (1.5) | 72 (4.3) |
| Overweight - Yes | 1,887 (39.3) | 796 (44.4) | 1,277 (44.8) | 868 (51.4) |
| **Household co-variates at 9m** |  |  |  |  |
| House owner (with or without mortgage) | 4,085 (85) | 1,403 (78.3) | 1,339 (47) | 578 (34.2) |
| *Family social class* |  |  |  |  |
| Professional/managerial | 3,471 (72.3) | 1,032 (57.6) | 672 (23.6) | 251 (14.9) |
| Other non-manual/skilled-manual | 1,066 (22.2) | 582 (32.5) | 1,195 (41.9) | 528 (31.3) |
| Semi-skilled/unskilled manual | 214 (4.5) | 130 (7.3) | 442 (15.5) | 290 (17.2) |
| Never worked at all | 53 (1.1) | 47 (2.6) | 542 (19) | 619 (36.7) |
| *Equivalised Household Income* |  |  |  |  |
| 4th-10th decile | 4,311 (89.7) | 1,578 (88.1) | 820 (28.8) | 351 (20.8) |
| 1st-3rd decile | 130 (2.7) | 97 (5.4) | 1,782 (62.5) | 1,201 (71.1) |
| *4 Category Household Type* |  |  |  |  |
| One parent + 1 child | 69 (1.4) | 74 (4.1) | 276 (9.7) | 171 (10.1) |
| One parent + >2 child | 46 (1) | 42 (2.3) | 339 (11.9) | 342 (20.3) |
| Two parents + 1 child | 2,054 (42.8) | 663 (37) | 700 (24.6) | 277 (16.4) |
| Two parents + >2 child | 2,635 (54.9) | 1,012 (56.5) | 1,536 (53.9) | 898 (53.2) |
| **Child outcomes at age 9** |  |  |  |  |
| Overweight | 581 (12.1) | 305 (17) | 451 (15.8) | 320 (19) |
| Longstanding illness | 710 (14.8) | 355 (19.8) | 441 (15.5) | 303 (18) |
| Total SDQ Difficulties<sup>3</sup> – Teacher | 334 (7) | 166 (9.3) | 302 (10.6) | 246 (14.6) |
| Total SDQ Difficulties<sup>3</sup> – Parent | 130 (2.7) | 107 (6) | 158 (5.5) | 147 (8.7) |
| **Mother outcomes at age 9** |  |  |  |  |
| Raised depression score (>17 score) | 168 (3.5) | 147 (8.2) | 182 (6.4) | 215 (12.7) |
| Longstanding illness | 481 (10) | 333 (18.6) | 364 (12.8) | 326 (19.3) |
| Overweight | 1,515 (31.5) | 702 (39.2) | 911 (32) | 608 (36) |

**Abbreviations:** PCG primary care giver; SDQ strengths and difficulties questionnaire.
<sup>1</sup> Vulnerable to Poverty 1: Material>Income; <sup>2</sup> Vulnerable to Poverty 2: Income > Material;
<sup>3</sup>SDQ >17 score indicates high risk.

### Primary analyses

#### Child outcomes

**Table 2** and **Figure 1, S5** shows the association of identified poverty trajectories and child outcomes at 9 years of age. In adjusted analysis, persistent poverty, compared to those never in poverty, was associated with higher odds (aOR adjusted odds ratio) of psychopathology reported by mother (aOR 2.06; 95% CI 1.42, 2.99) and teacher (aOR 1.99 95% CI 1.50, 2.64), and higher odds of physical health problems (being overweight (aOR 1.70 95% CI 1.24, 2.16) or higher odds of having a longstanding health condition (aOR 1.51 95% 1.20, 1.91). Vulnerable to poverty group 2 in comparison to never in poverty had lower odds of mother reported (aOR 1.51, 95% 1.07, 2.12) and higher odds of teacher reported (aOR 1.47 95% 1.15, 1.88) psychopathology.

**Table 2:**
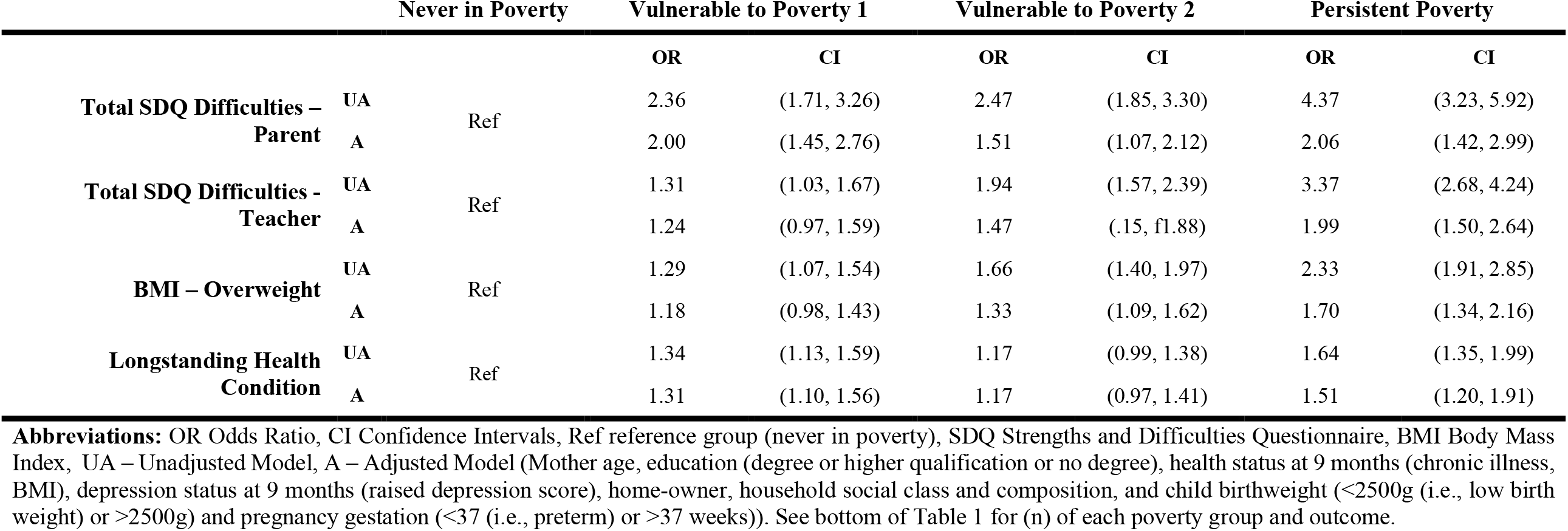
Association of predicted poverty trajectories from 9 months to 9 years and child outcomes at 9 years using the Irish Growing Up in Ireland – Infant Cohort.

**Figure 1:**
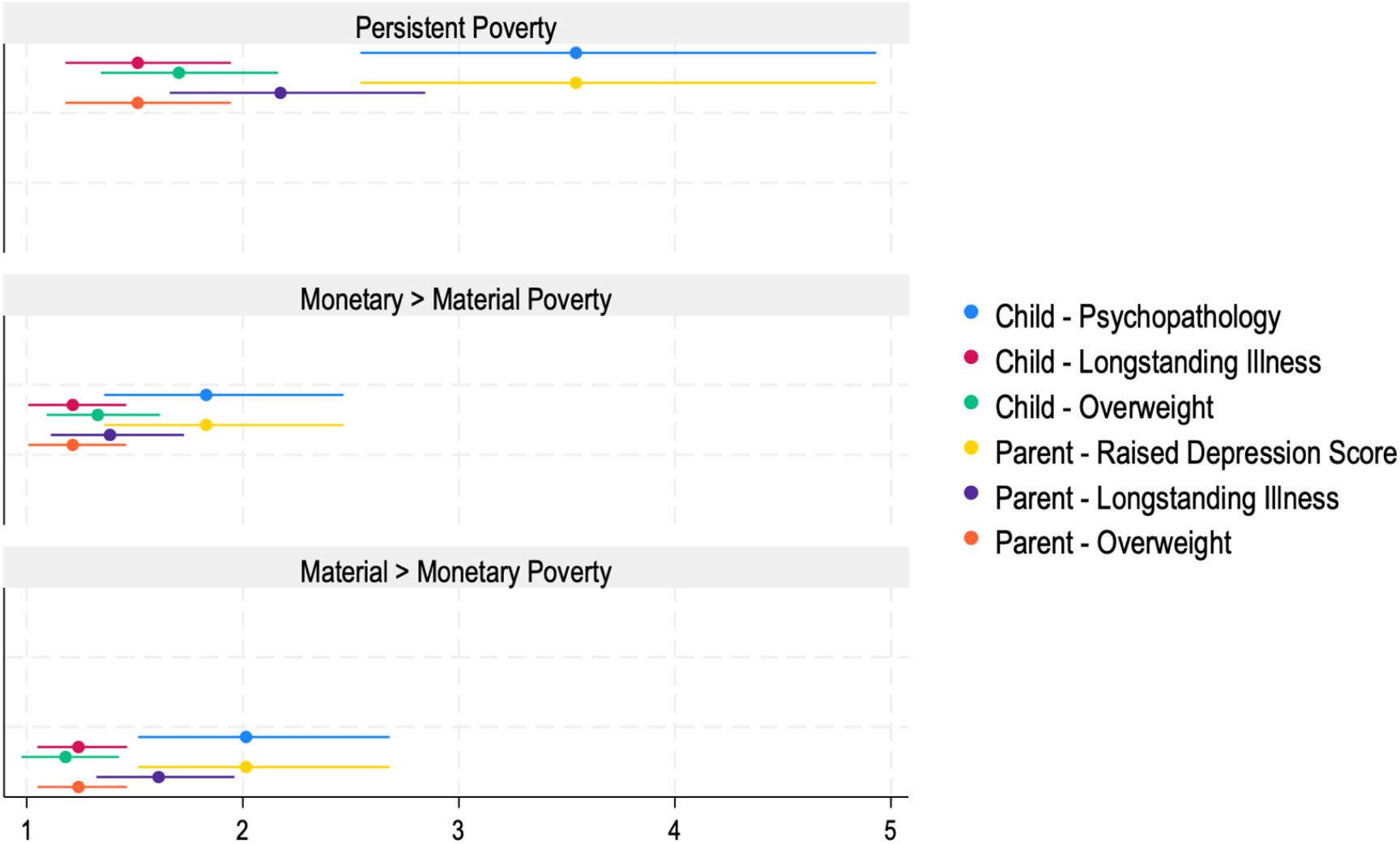
Association of poverty trajectories (persistent poverty, monetary > material poverty, and material > monetary poverty) and child outcomes at 9 years of age (psychopathology measured by total difficulties by strengths and difficulties questionnaire, having a longstanding illness, and being overweight) and parent (mother) outcomes after 9 years (raised depression score, having a longstanding illness and being overweight). Child model adjusted for mother age, education (degree or higher qualification or no degree), health status at 9 months (chronic illness, BMI), depression status at 9 months (raised depression score), home-ownership, household social class and composition, and child birthweight (<2500g (i.e., low birth weight) or >2500g) and pregnancy gestation (<37 (i.e., preterm) or >37 weeks). Parent (mother) model adjusted for mother age, education (degree or higher qualification or no degree), health status when child 9 months (chronic illness, BMI), depression status when child 9 months (raised depression score), home-ownership, household social class and composition. See supplementary material (**Figure S5, Figure S6**) for expanded figure including unadjusted and adjusted models.

#### Mother outcomes

**Table 3** and **Figure 1, S6** shows the association of identified poverty trajectories and mother outcomes when the child is 9 years of age. In adjusted analysis, persistent poverty, compared to those never in poverty, was associated with higher odds of raised depression scores (Aor 3.54 95% CI 2.54, 4.94), of being overweight (aOR 1.49 95% CI 1.16, 1.92) and of having a longstanding health condition (aOR 2.13 95% CI 1.63, 2.79). Vulnerable to poverty group 1 in comparison to never in poverty had higher odds of raised depression scores (aOR 2.01 95% CI 1.51 2.67), of being overweight (aOR 1.23 95% CI 1.5, 2.17) and of having a longstanding health condition (aOR 1.60 95% CI 1.31, 1.95).

**Table 3:**
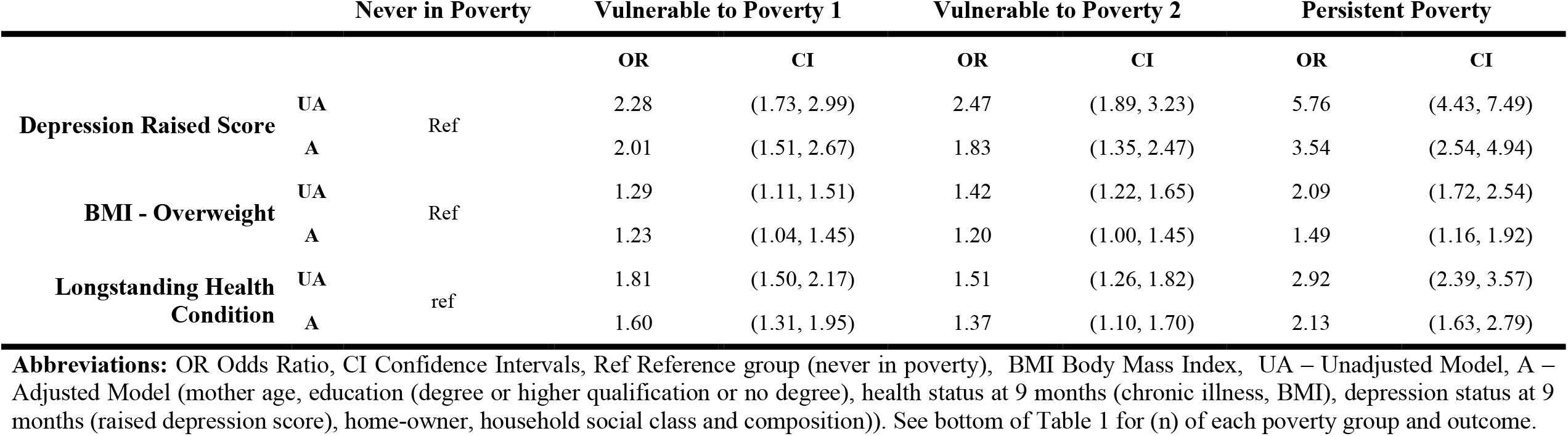
Association of predicted poverty trajectories mother outcomes after 9 years using the Irish Growing Up in Ireland – Infant Cohort.

### Subgroup analyses

#### Attendance at an extra resourced school (DEIS) in an area of deprivation with child and mother outcomes

Children identified as being in persistent poverty and attend a DEIS school (extra resourced school (DEIS) in an area of deprivation) had attenuation of the odds of psychopathology parent reported (aOR 1.66 95% CI 0.72, 3.81) or teacher reported (aOR 1.78 95% CI 0.91, 3.46), odds of being overweight (aOR 1.38 95% CI 0.80, 2.41) or having a longstanding illness (aOR 0.95 95% CI 0.56, 1.62) compared to child that attend a non-DEIS school (psychopathology parent reported (aOR 1.76 95% CI 1.10, 2.83) or teacher reported (aOR 1.93 95% CI 1.38, 2.70), being overweight (aOR 1.78 95% CI 1.33, 2.36), and having a longstanding illness (aOR 1.67 95% CI 1.26, 2.21) (**Table S7**). Mothers in persistent poverty and their child attending a DEIS school had attenuation of the odds of having raised depression scores (aOR 2.60 95% CI 1.20, 5.66) and being overweight (aOR 1.19 95% CI 0.66, 2.13), in comparison to their child attending a non-DEIS school (raised depression scores (aOR 3.53 95% CI 2.39, 5.21), and being overweight (aOR 1.45 95% CI 1.09, 1.94) (**Table S8**).

#### Child sex with child and mother outcomes

Child girls identified as living in a persistent poverty trajectory and had higher odds of psychopathology scores (aOR 2.22 95% CI 1.2, 4.07) compared to boys (aOR 1.97 95% CI 1.23, 3.15), and there was no sex interaction (p=0.3479) (**Table S9**). Contrastingly, child boys identified as living in a persistent poverty trajectory had higher odds of having a longstanding illness (aOR 1.76 95% CI 1.29, 2.41) compared to girls (aOR 1.18 95% CI 0.82, 1.69), and there was no sex specific interaction (p=0.2408) (**Table S9**). Mothers with child who is a boy living in persistent poverty trajectory had higher odds of a raised depression scores (aOR 4.15 95% CI 2.57, 6.69) and lower odds of being overweight (aOR 1.33 95% CI 0.95, 1.87), compared to mothers with a child who is a girl (raised depression scores (aOR 3.02 95% CI 1.88, 4.83), being overweight (aOR 1.71 95% CI 1.21, 2.43) (**Table S10**).

### Sensitivity analyses

The sensitivity analysis using imputed data showed similar patterns of associations as the main analysis (**Table S11, Table S12**). We continued to report odds ratio, as the incidence risk ratio of child and mother outcomes were marginally lower than reported odds ratios (**Table S13, Table S14**).

## Discussion

To our knowledge, this is the first study to assess clustering of trajectories of multi-dimensional aspects of poverty from 9 months to 9 years and their association with child and mother outcomes using a large prospective representative Irish sample. We show that children who experience some trajectory of poverty experience, have higher odds of being overweight, of having a longstanding illness and of having higher psychopathology scores by age 9 years. Similarly, mothers that experience some trajectory of poverty experience had higher odds of depression, being overweight and having a longstanding illness. Over 15% of children in our study had a persistent poverty trajectory from 9 months to 9 years of age. Positively, children living in a household experiencing poverty who attended a DEIS school (extra resourced school in an area of deprivation) had lower odds of child and mother mental and physical health problems in comparison to children who did not attend a DEIS school.

We identified four distinct trajectories of poverty during childhood and found that any poverty trajectory was associated with increased risk of poor childhood outcome at 9 years of age. Similar to previous research, we showed that the persistent poverty trajectory is associated with worse outcomes in mid-childhood (Adjei et al., 2022; Lai et al., 2019). We illustrate a lower odds ratio in our associations than previously published studies. This may be related to country differences and due to our approach to adjusting for potential confounders. Uniquely, we highlight that material poverty without monetary poverty is still associated with worse outcomes and that the additive monetary poverty appears to be cumulative with a higher odds of poor mental and physical outcomes.

The most important finding of our study is that children and mothers experiencing any poverty trajectory have poorer outcomes, with persistent poverty having the worse outcomes in comparison to households never in poverty. From early childhood into adolescence is a critical time period for development, especially the development of mental disorders that may peak during adolescence. Hence the importance of focused policies that aim to intervene at not just the individual level but system level (family, school, community). Strategies that target health outcomes (e.g., mental health) without addressing child poverty will be challenged due to increased need for services arising out of rising incidence of poverty. In contrast, strategies that intervene at a system level (i.e., school, community) to target or reach the individual level may have broader positive effects during the life course for many health outcomes.

There must be an acceptance that measuring poverty should be carefully considered dependent on country, regional area, social services availability and family factors. Defining poverty and using the correct measurement of poverty during the life course is subject to ongoing debate. This means that there are no collectively agreed standard measures to facilitate comparability of findings. As authors, we argue for a multi-dimensional approach to measuring poverty in research that encompasses the material, monetary and subjective dimensions.

This study is the first to identify clustering of poverty trajectories across the early life course using a multi-dimensional poverty approach. While trajectories of a child’s outcomes and poverty adversity may be connected during the life course, our approach takes into consideration a child’s poverty adversity (i.e., criterial developmental periods) over time. Poverty is an adversity, and emerging evidence suggests that multiple-adversities may be synergistic (Adjei et al., 2022). As such, a public policy such as DEIS may be supporting households in poverty with funded support in education and have an unmeasured association on other adversities (e.g., parental depression). Our study shows that mothers in any poverty trajectory have lower odds of depression, and of being overweight if their child attends a DEIS school. Moreover, in our opinion public policy interventions to improve child outcomes, which do not address poverty may have a reduced impact. By doing same, there should be the development of a more integrated social and health policy approach to addressing poverty and reducing socio-economic inequality.

A main strength of the study is that we used a contemporary national representative Irish cohort, that has generalisable properties to the Irish population and policy. We used a rigorous modelling technique to identify poverty trajectories that allowed clustering of at risk poverty groups over time. Moreover this technique maximised the included sample size as it includes participants that have available data at a minimum of one time point. Our results were also similar when we did multiple imputation to account for missing data on outcomes.

Regardless of these strengths, this study must be understood with reference to the following limitations. First, there may be bias in the class estimates with outcomes and reduced standard errors. This is due to the group based trajectory modelling approach that assumes classes are independent and that all potential participants chart the same trajectory. Practically, some participants would vary in trajectory and this cannot be estimated with this approach. Despite this, it is unlikely to influence the results as we were interested in the class membership and their association with outcomes rather than distinct shape of class of trajectory over time.

Second, the SDQ and CES-D outcomes are dependent on the subjective evaluation by a caregiver or teacher. Both questionnaires have good correlation identifying at risk participants (Muris et al., 2003; Van Roy et al., 2008). Teacher and parent SDQ have good inter-rater agreement, which is helpful if a parent with depression completed both the CES-D and SDQ. These measures do not provide a clinical diagnosis. Third, paternal or secondary care giver outcomes were not investigated in this study and the authors are of the view this is a separate distinct study due to changes in secondary care giver status, and the need for a separate conceptualising approach. Fourth, loss to follow in longitudinal studies may be higher in families with adversity and mental illness. Our approach in addressing missing data maximises participants that did contribute to the cohort at least once. Fifth, while our findings are similar to previous studies, we cannot state that particular poverty trajectories are causal of outcomes, and that there may be potential unmeasured confounding factors not included (e.g., genetic risk factor). Sixth, while attendance at a DEIS school was associated with lower odds ratios of outcomes, due to unmeasured factors (e.g., quality of teacher experience), we cannot state this is causal, yet it warrants further large-scale studies to determine if DEIS school attendance with a randomized long-term follow up has wider societal benefit.

## Conclusion

In this large prospective nationally representative cohort, we demonstrated that predicted poverty trajectories are associated with adverse health outcomes at early childhood for children 9 (age 9) and their mothers. Moreover, policy initiatives to address educational disadvantage appeared to mitigate the effect of child poverty on both mother and child outcomes and may have wider positive public health consequences with improved mental and physical wellbeing during the life course. This work should be replicated in different cohorts. Further work is required to better understand if these predicted poverty trajectories allow us to identify distinct subgroups of the population that require additional support or intervention, and whether further policy development and interventions (e.g., DEIS) can be used to ameliorate the negative effects of the growing problem that is child poverty.

## Data Availability

Data may be accessed by applying to the Central Statistics Office of Ireland.

## Acknowledgements

The authors received approval from the Central Statistics Office to use the AMF Infant Cohort dataset. The GUI study received ethical approval from the Research Ethics Committee within the Irish Department of Health and Children, Ireland. All participants gave informed consent to enrol in the GUI study. This study is in accordance with the ethical standards as per the Declaration of Helsinki (1964) and subsequent amendments. The present analyses did not require additional ethical approval.

