## Supplementary Material for "Poverty trajectories and child and mother wellbeing outcomes in Ireland: findings from an Irish prospective cohort"

**Table S1:** Description of household poverty exposure variables
**Table S2:** Material deprivation index breakdown of questions using the Irish Growing Up in Ireland - Infant Cohort
**Table S3**: Centre for epidemiological studies depression scale (8-item) (CESD-8)
**Table S4:** Model Selection Results
**Table S5:** Model Adequacy Results
**Table S6:** Description of group based trajectory subgroups and key variables by age using the Irish Growing Up in Ireland Study – Infant Cohort.
**Table S7**: Association of poverty trajectories from 9 months to 9 years and adjusted child outcomes at 9 years, stratified by DEIS school attendance and interaction analysis using the Irish Growing Up in Ireland Study - Infant Cohort
**Table S8:** Association of poverty trajectories from 9 months to 9 years and mother outcomes at 9 years, stratified by DEIS school attendance and interaction analysis using the Irish Growing Up in Ireland Study - Infant Cohort
**Table S9:** Association of poverty trajectories from 9 months to 9 years and child outcomes at 9 years, stratified by child sex and interaction analysis using the Irish Growing Up in Ireland Study - Infant Cohort
**Table S10:** Association of poverty trajectories from 9 months to 9 years and mother outcomes at 9 years, stratified by child sex and interaction analysis using the Irish Growing Up in Ireland Study - Infant Cohort
**Table S11**: Imputed data: association of predicted poverty trajectories from 9 months to 9 years and adjusted child outcomes at 9 years, using the Irish Growing Up in Ireland Study - Infant Cohort.
**Table S12:** Imputed data: association of predicted poverty trajectories from 9 months to 9 years and mother outcomes at 9 years, using the Irish Growing Up in Ireland Study - Infant Cohort.
**Table S13:** Sensitivity analysis – reporting incidence of risk ratio in the association of predicted poverty trajectories from 9 months to 9 years and child outcomes at 9 years using the Irish Growing Up in Ireland – Infant Cohort
**Table S14:** Sensitivity analysis – reporting incidence of risk ratio in the association of predicted poverty trajectories mother outcomes after 9 years using the Irish Growing Up in Ireland – Infant Cohort.
**Figure S1:** Group based trajectory of poverty using the Irish Growing Up in Ireland - Infant Cohort
**Figure S2:** Group based trajectory of poverty of lowest third decile income, ease to meet bills (difficult or very difficult), and number of material deprivations (0-6) into four trajectories: (A) never poverty (43.1%), (B) vulnerable to poverty: material/perceived>monetary (16.1%), (C) vulnerable to poverty: monetary>material/perceived (25.6%) and (D) persistent poverty (15.2%) using the Irish Growing Up in Ireland - Infant Cohort.
**Figure S3:** Directed Acyclic Graph (DAG) of association between poverty trajectory and child outcomes using the Irish Growing Up in Ireland - Infant Cohort
**Figure S4:** Directed Acyclic Graph (DAG) of association between poverty trajectory and mother outcomes using the Irish Growing Up in Ireland - Infant Cohort

**Figure S5:** Associations of poverty trajectories (never in poverty, vulnerable to material poverty (group 1), vulnerable to material and income poverty (group 2) and persistent poverty) and child outcomes (total strengths and difficulties (SDQ) scores by mother and by teacher, longstanding illness, and overweight) at age 9 in the Irish Growing Up in Ireland Study – Infant cohort. Models were unadjusted (UA) and adjusted (A) for mother age, education (degree or higher qualification or no degree), health status at 9 months (chronic illness, BMI), depression status at 9 months (raised depression score), home-owner, household social class and composition, and child birthweight (<2500g (i.e., low birth weight) or >2500g) and pregnancy gestation (<37 (i.e., preterm) or >37 weeks).

**Figure S6:** Associations of poverty trajectories (never in poverty, vulnerable to material poverty (group 1), vulnerable to material and income poverty (group 2) and persistent poverty) and mother outcomes (raised depression score, longstanding illness, and overweight) at age 9 in the Irish Growing Up in Ireland Study – Infant cohort. Models were unadjusted (UA) and adjusted (A) for PCG age, education (degree or higher qualification or no degree), health status at 9 months (chronic illness, BMI), depression status at 9 months (raised depression score), home-owner, household social class and composition.

**Supplementary material**

**Tables**

**Table S1:** Description of household poverty exposure variables

| | |
|---|---|
| **Monetary poverty** | The monetary poverty variable obtained by parental interview included the three lowest deciles of household equivalised income (computed as three binary dummy variables). |
| **Subjective sense of poverty** | For the subjective sense of poverty, participants were asked "concerning your household's total monthly or weekly income, with which degree of ease or difficulty is the household able to make ends meet?". If a participant responded with great difficulty or difficulty, they were coded as having a high subjective sense of poverty. |
| **Material poverty** | For material poverty, participants were asked questions about household deprivation and could indicate three options in response to having a household item 'yes', or 'no', 'can't afford or no, other reason'. See **Table S1** for material deprivation index survey questions. A total score was obtained of items to compute a material deprivation index. For group based trajectory analysis, material deprivation was grouped into six categories of scores (0, 1, 2, 3, 4, 5, 6+ score). |

**Table S2:** Material deprivation index breakdown of questions using the Irish Growing Up in Ireland - Infant Cohort

| | |
|---|---|
| For the following items could you indicate whether or not your household, has the item and, if not, if it is because you couldn't afford it or for another reason? Three options in response: (i) Yes (ii) No, cannot afford (iii) No, other reason | |
| **1** | Does your household eat meals with meat, chicken, fish (or vegetarian equivalent) at least every second day? |
| **2** | Does your household have a roast joint (or its equivalent) at least once a week? |
| **3** | Do household members buy new rather than second-hand clothes? |
| **4** | Does each household member possess a warm waterproof coat? |
| **5** | Does each household member possess two pairs of strong shoes? |
| **6** | Does the household replace any worn out furniture? |
| **7** | Does the household keep the home adequately warm? |
| **8** | Does the household have family or friends for a drink or meal once a month? |
| **9** | Does the household buy presents for family or friends at least once a year? |

Where household 'cannot afford' two or more items from the above list is used as an indication of household deprivation.

**Table S3**: Centre for epidemiological studies depression scale (8-item) (CESD-8)

Listed on this card are 8 statements about some of the ways you may have felt or behaved. Please indicate how often you have felt this way during the past week. Four options in response to each statement below:
Rarely or none of the time (less than 1 day) (score 0)
Some or a little of the time (1-2 days) (score 1)
Occasionally or a moderate amount of the time (3-4 days) (score 2)
Most or all of the time (5-7 days) (score 3)

| | |
|---|---|
| **1** | I felt I could not shake off the blues even with help from my family or friends. |
| **2** | I felt depressed. |
| **3** | I thought my life had been a failure. |
| **4** | I felt fearful. |
| **5** | My sleep was restless. |
| **6** | I felt lonely. |
| **7** | I had crying spells. |
| **8** | I felt sad. |

A composite score is calculated (0-24). Respondents are categorised according to the recommended criterion for depression, with composite scores of 7 being classified as depressed and scores

T**able S4:** Model Selection Results

| | BIC | AIC | Entropy |
|---|---|---|---|
| **One Group** | -97446.6463 | -97402.83316 | Ref |
| **Two Groups** | -80828.81849 | -80737.54112 | 0.9 |
| **Three Groups** | -78094.4508 | -77955.70919 | 0.8 |
| **Four Groups** | -76962.98108 | -76776.77524 | 0.8 |
| **Five Groups** | -76310.25519 | -76076.58511 | 0.8 |
| **Six Groups** | -76225.45686 | -75944.32254 | 0.7 |

**Abbreviations:** BIC – Bayesian information criterion; AIC – Akaike information criterion

**Table S5:** Model Adequacy Results

| Trajectory group | n (%) | AvePP | OCC |
|---|---|---|---|
| Never in Poverty | 4804 (39.5) | 0.88 | 10.09 |
| Vulnerable to Poverty 1 | 1791 (18.9) | 0.87 | 35.22 |
| Vulnerable to Poverty 2 | 2851 (25.7) | 0.87 | 19.84 |
| Persistent Poverty | 1688 (15.9) | 0.91 | 57.12 |

**Abbreviations:** AvePP – average posterior probability; OCC – odds of correct classification. A good fit model (assessment of adequacy) was determined by subgroup membership of AvePP>0.70 and OCC>5.

**Table S6:** Description of group based trajectory subgroups and key variables by age using the Irish Growing Up in Ireland Study – Infant Cohort.

| | Never in Poverty | Vulnerable to Poverty 1 | Vulnerable to Poverty 2 | Persistent Poverty |
|---|---|---|---|---|
| | **(n=4,804)** | **(n=1,791)** | **(n=2,851)** | **(n=1,688)** |
| **Lowest-3rd decile equivalised household income** | | | | |
| Age 9 months - Yes | 130 ( 2.9) | 97 ( 5.8) | 1,782 (68.5) | 1,201 (77.4) |
| Age 3 years – Yes | 59 ( 1.5) | 99 ( 6.2) | 1,486 (64.9) | 1,031 (74.9) |
| Age 5 years – Yes | 64 ( 1.7) | 82 ( 5.3) | 1,323 (63.1) | 947 (76.1) |
| Age 9 years - Yes | 127 ( 3.8) | 112 ( 8.4) | 901 (53.7) | 678 (72.6) |
| **Someone in household receiving welfare** | | | | |
| Age 9 months - Yes | 464 ( 9.7) | 310 (17.3) | 934 (32.8) | 675 (40) |
| Age 3 years – Yes | 650 (15.4) | 508 (30) | 1,409 (57.6) | 1,057 (73.5) |
| Age 5 years – Yes | 537 (13.8) | 466 (29.1) | 1,265 (57.1) | 944 (73.5) |
| Age 9 years - Yes | 408 (11.3) | 314 (21.4) | 820 (43.6) | 669 (63.1) |
| **Difficulty to make ends meet [1]** | | | | |
| Age 9 months - Yes | 108 (2.2) | 362 (20.2) | 406 (14.2) | 1,033 (61.2) |
| Age 3 years – Yes | 102 (2.4) | 507 (29.9) | 414 (16.9) | 912 (63.4) |
| Age 5 years – Yes | 158 (4.1) | 652 (40.7) | 430 (19.4) | 928 (72.2) |
| Age 9 years - Yes | <30 | 57 ( 3.9) | <30 | 176 (16.6) |
| **Total material deprivation Score[2]** | | | | |
| Age 9 months >2 | 80 (1.7) | 193 (10.8) | 368 (12.9) | 918 (54.4) |
| Age 3 years >2 | 0 (0) | 149 ( 8.8) | 73 (3.0) | 719 (50) |
| Age 5 years >2 | 0 (0) | 452 (28.2) | 203 (9.2) | 924 (71.9) |
| Age 9 years >2 | 0 (0) | 119 ( 8.1) | 40 (2.1) | 419 (39.5) |

[1] Difficulty or very difficult at paying bills.
[2] Total Deprivation Score out of 11, illustrated as >2 which indicates material deprivation risk.

**Table S7**: Association of poverty trajectories from 9 months to 9 years and adjusted child outcomes at 9 years, stratified by DEIS school attendance and interaction analysis using the Irish Growing Up in Ireland Study - Infant Cohort

| | No DEIS | | | | Yes DEIS | | | | Interaction |
|---|---|---|---|---|---|---|---|---|---|
| | **(n)** | **OR** | **LCI** | **UCI** | **(n)** | **OR** | **LCI** | **UCI** | **p-value** |
| **Never in poverty (reference)** | | | | | | | | | |
| **Vulnerable to poverty 1** | | | | | | | | | |
| Raised Parent Total SDQ score | 75 | 1.83 | 1.26 | 2.66 | 18 | 1.84 | 0.77 | 4.39 | 0.4812 |
| Raised Teacher Total SDQ score | 138 | 1.30 | 1.00 | 1.71 | 21 | 0.97 | 0.47 | 2.02 | 0.5213 |
| Overweight | 44 | 1.14 | 0.92 | 1.42 | 12 | 1.23 | 0.71 | 2.12 | 0.7536 |
| Longstanding illness | 256 | 1.25 | 1.02 | 1.53 | 48 | 0.83 | 0.49 | 1.41 | 0.6688 |
| **Vulnerable to poverty 2** | | | | | | | | | |
| Raised Parent Total SDQ score | 103 | 1.67 | 1.11 | 2.49 | 40 | 1.08 | 0.48 | 2.42 | 0.2852 |
| Raised Teacher Total SDQ score | 208 | 1.50 | 1.13 | 1.98 | 75 | 1.33 | 0.72 | 2.46 | 0.9645 |
| Overweight | 66 | 1.28 | 1.02 | 1.62 | 32 | 1.54 | 0.96 | 2.47 | 0.4069 |
| Longstanding illness | 287 | 1.26 | 1.01 | 1.56 | 103 | 0.73 | 0.44 | 1.19 | 0.4064 |
| **Persistent poverty** | | | | | | | | | |
| Raised Parent Total SDQ score | 82 | 1.76 | 1.10 | 2.83 | 48 | 1.66 | 0.72 | 3.81 | 0.7441 |
| Raised Teacher Total SDQ score | 161 | 1.93 | 1.38 | 2.70 | 78 | 1.78 | 0.91 | 3.46 | 0.8457 |
| Overweight | 51 | 1.78 | 1.33 | 2.36 | 32 | 1.38 | 0.80 | 2.41 | 0.2041 |
| Longstanding illness | 192 | 1.67 | 1.26 | 2.21 | 81 | 0.95 | 0.56 | 1.62 | 0.9785 |

**Abbreviations:** OR odds ratio, LCI lower confidence interval (95), UCI upper confidence interval (95), SDQ strengths and difficulties questionnaire.
**Adjusted** by mother age, education (degree or higher qualification or no degree), health status at 9 months (chronic illness, BMI), depression status at 9 months (raised depression score), home-owner, household social class and composition, and child birthweight (<2500g (i.e., low birth weight) or >2500g) and pregnancy gestation (<37 (i.e., preterm) or >37 weeks). See bottom of Table 1 for (n) of each poverty group and outcome.

**Table S8:** Association of poverty trajectories from 9 months to 9 years and mother outcomes at 9 years, stratified by DEIS school attendance and interaction analysis using the Irish Growing Up in Ireland Study - Infant Cohort

| | No DEIS | | | | Yes DEIS | | | | Interaction |
|---|---|---|---|---|---|---|---|---|---|
| | **(n)** | **OR** | **LCI** | **UCI** | **(n)** | **OR** | **LCI** | **UCI** | **p-value** |
| **Never in poverty (reference)** | | | | | | | | | |
| **Vulnerable to poverty 1** | | | | | | | | | |
| Raised Depression | 114 | 1.90 | 1.38 | 2.61 | 23 | 1.62 | 0.77 | 3.41 | 0.5293 |
| Overweight | 185 | 1.27 | 1.05 | 1.52 | 48 | 1.06 | 0.63 | 1.77 | 0.1857 |
| Longstanding illness | 253 | 1.77 | 1.42 | 2.21 | 43 | 1.06 | 0.59 | 1.89 | 0.4488 |
| **Vulnerable to poverty 2** | | | | | | | | | |
| Raised Depression | 123 | 1.97 | 1.41 | 2.76 | 33 | 0.84 | 0.40 | 1.79 | 0.0657 |
| Overweight | 243 | 1.19 | 0.97 | 1.47 | 102 | 0.93 | 0.57 | 1.52 | 0.2475 |
| Longstanding illness | 228 | 1.55 | 1.21 | 1.98 | 91 | 1.19 | 0.66 | 2.14 | 0.1926 |
| **Persistent poverty** | | | | | | | | | |
| Raised Depression | 128 | 3.53 | 2.39 | 5.21 | 70 | 2.60 | 1.20 | 5.66 | 0.1152 |
| Overweight | 163 | 1.45 | 1.09 | 1.94 | 100 | 1.19 | 0.66 | 2.13 | 0.9110 |
| Longstanding illness | 187 | 2.20 | 1.60 | 3.02 | 102 | 2.26 | 1.17 | 4.35 | 0.1041 |

**Abbreviations:** OR odds ratio, LCI lower confidence interval (95), UCI upper confidence interval (95), SDQ strengths and difficulties questionnaire.
**Adjusted** by mother age, education (degree or higher qualification or no degree), health status at 9 months (chronic illness, BMI), depression status at 9 months (raised depression score), home-owner, household social class and composition. See bottom of Table 1 for (n) of each poverty group and outcome.

**Table S9:** Association of poverty trajectories from 9 months to 9 years and child outcomes at 9 years, stratified by child sex and interaction analysis using the Irish Growing Up in Ireland Study - Infant Cohort

| | Male | | | | Female | | | | Interaction |
|---|---|---|---|---|---|---|---|---|---|
| | **(n)** | **OR** | **LCI** | **UCI** | **(n)** | **OR** | **LCI** | **UCI** | **p-value** |
| **Never in poverty (reference)** | | | | | | | | | |
| **Vulnerable to poverty 1** | | | | | | | | | |
| Raised Parent Total SDQ score | 54 | 1.48 | 0.96 | 2.28 | 53 | 3.15 | 1.89 | 5.23 | 0.0203 |
| Raised Teacher Total SDQ score | 108 | 1.11 | 0.82 | 1.50 | 58 | 1.67 | 1.08 | 2.58 | 0.8752 |
| Overweight | 33 | 1.07 | 0.80 | 1.44 | 29 | 1.26 | 0.98 | 1.62 | 0.1217 |
| Longstanding illness | 200 | 1.35 | 1.06 | 1.71 | 155 | 1.29 | 0.99 | 1.68 | 0.5176 |
| **Vulnerable to poverty 2** | | | | | | | | | |
| Raised Parent Total SDQ score | 93 | 1.41 | 0.93 | 2.14 | 65 | 1.70 | 0.95 | 3.04 | 0.7815 |
| Raised Teacher Total SDQ score | 191 | 1.27 | 0.94 | 1.72 | 111 | 2.17 | 1.42 | 3.30 | 0.3456 |
| Overweight | 60 | 1.50 | 1.13 | 1.99 | 49 | 1.18 | 0.90 | 1.56 | 0.0894 |
| Longstanding illness | 252 | 1.29 | 1.00 | 1.66 | 189 | 1.00 | 0.76 | 1.33 | 0.8258 |
| **Persistent poverty** | | | | | | | | | |
| Raised Parent Total SDQ score | 101 | 1.97 | 1.23 | 3.15 | 46 | 2.22 | 1.21 | 4.07 | 0.3479 |
| Raised Teacher Total SDQ score | 153 | 1.62 | 1.13 | 2.33 | 93 | 2.94 | 1.86 | 4.65 | 0.2676 |
| Overweight | 46 | 1.79 | 1.27 | 2.53 | 48 | 1.66 | 1.20 | 2.32 | 0.6329 |
| Longstanding illness | 196 | 1.76 | 1.29 | 2.41 | 107 | 1.18 | 0.82 | 1.69 | 0.2408 |

**Abbreviations:** OR odds ratio, LCI lower confidence interval (95), UCI upper confidence interval (95), SDQ strengths and difficulties questionnaire.
**Adjusted** by mother age, education (degree or higher qualification or no degree), health status at 9 months (chronic illness, BMI), depression status at 9 months (raised depression score), home-owner, household social class and composition, and child birthweight (<2500g (i.e., low birth weight) or >2500g) and pregnancy gestation (<37 (i.e., preterm) or >37 weeks). See bottom of Table 1 for (n) of each poverty group and outcome.

**Table S10:** Association of poverty trajectories from 9 months to 9 years and mother outcomes at 9 years, stratified by child sex and interaction analysis using the Irish Growing Up in Ireland Study - Infant Cohort

| | Male | | | | Female | | | | Interaction |
|---|---|---|---|---|---|---|---|---|---|
| | (n) | OR | LCI | UCI | (n) | OR | LCI | UCI | p-value |
| **Never in poverty (reference)** | | | | | | | | | |
| **Vulnerable to poverty 1** | | | | | | | | | |
| Raised Depression | 76 | 2.37 | 1.57 | 3.56 | 71 | 1.71 | 1.15 | 2.54 | 0.4554 |
| Overweight | 118 | 1.02 | 0.80 | 1.30 | 139 | 1.47 | 1.16 | 1.85 | 0.0239 |
| Longstanding illness | 167 | 1.63 | 1.23 | 2.16 | 166 | 1.52 | 1.14 | 2.01 | 0.8604 |
| **Vulnerable to poverty 2** | | | | | | | | | |
| Raised Depression | 86 | 2.05 | 1.33 | 3.15 | 96 | 1.65 | 1.09 | 2.52 | 0.7609 |
| Overweight | 195 | 1.28 | 0.99 | 1.65 | 187 | 1.13 | 0.88 | 1.45 | 0.1684 |
| Longstanding illness | 182 | 1.47 | 1.05 | 2.04 | 182 | 1.25 | 0.93 | 1.67 | 0.6520 |
| **Persistent poverty** | | | | | | | | | |
| Raised Depression | 119 | 4.15 | 2.57 | 6.69 | 96 | 3.02 | 1.88 | 4.83 | 0.6532 |
| Overweight | 145 | 1.33 | 0.95 | 1.87 | 150 | 1.71 | 1.21 | 2.43 | 0.4421 |
| Longstanding illness | 168 | 2.17 | 1.48 | 3.20 | 158 | 2.06 | 1.42 | 2.98 | 0.4831 |

**Abbreviations:** OR odds ratio, LCI lower confidence interval (95), UCI upper confidence interval (95), SDQ strengths and difficulties questionnaire.
**Adjusted** by mother age, education (degree or higher qualification or no degree), health status at 9 months (chronic illness, BMI), depression status at 9 months (raised depression score), home-owner, household social class and composition. See bottom of Table 1 for (n) of each poverty group and outcome.

**Table S11**: Imputed data: association of predicted poverty trajectories from 9 months to 9 years and adjusted child outcomes at 9 years, using the Irish Growing Up in Ireland Study - Infant Cohort.

| | Unadjusted | | | Adjusted | | |
|---|---|---|---|---|---|---|
| | **OR** | **LCI** | **UCI** | **OR** | **LCI** | **UCI** |
| **Never in poverty (reference)** | | | | | | |
| **Vulnerable to poverty 1** | | | | | | |
| Raised Parent Total SDQ score | 2.09 | 1.63 | 2.68 | 1.83 | 1.43 | 2.36 |
| Raised Teacher Total SDQ score | 1.29 | 1.07 | 1.57 | 1.19 | 0.97 | 1.45 |
| Overweight | 1.36 | 1.16 | 1.58 | 1.22 | 1.04 | 1.44 |
| Longstanding illness | 1.33 | 1.14 | 1.55 | 1.29 | 1.10 | 1.51 |
| **Vulnerable to poverty 2** | | | | | | |
| Raised Parent Total SDQ score | 2.55 | 2.03 | 3.21 | 1.69 | 1.31 | 2.18 |
| Raised Teacher Total SDQ score | 1.97 | 1.69 | 2.30 | 1.44 | 1.18 | 1.74 |
| Overweight | 1.69 | 1.47 | 1.95 | 1.24 | 1.03 | 1.48 |
| Longstanding illness | 1.26 | 1.12 | 1.43 | 1.25 | 1.08 | 1.44 |
| **Persistent poverty** | | | | | | |
| Raised Parent Total SDQ score | 4.29 | 3.39 | 5.44 | 2.35 | 1.76 | 3.14 |
| Raised Teacher Total SDQ score | 3.23 | 2.74 | 3.81 | 2.00 | 1.62 | 2.45 |
| Overweight | 2.33 | 2.01 | 2.72 | 1.52 | 1.22 | 1.89 |
| Longstanding illness | 1.65 | 1.40 | 1.96 | 1.51 | 1.24 | 1.83 |

**Abbreviations:** OR odds ratio, LCI lower confidence interval (95), UCI upper confidence interval (95), SDQ strengths and difficulties questionnaire.
**Adjusted** by mother age, education (degree or higher qualification or no degree), health status at 9 months (chronic illness, BMI), depression status at 9 months (raised depression score), home-owner, household social class and composition, and child birthweight (<2500g (i.e., low birth weight) or >2500g) and pregnancy gestation (<37 (i.e., preterm) or >37 weeks). Sensitivity analysis using multiple imputation, (20 draws). See bottom of Table 1 for (n) of each poverty group and outcome.

**Table S12:** Imputed data: association of predicted poverty trajectories from 9 months to 9 years and mother outcomes at 9 years, using the Irish Growing Up in Ireland Study - Infant Cohort.

| | | **Unadjusted** | | | **Adjusted** | |
|---|---|---|---|---|---|---|
| | **OR** | **LCI** | **UCI** | **OR** | **LCI** | **UCI** |
| **Never in poverty (reference)** | | | | | | |
| **Vulnerable to poverty 1** | | | | | | |
| Raised Depression | 2.27 | 1.81 | 2.84 | 2.01 | 1.59 | 2.53 |
| Overweight | 1.37 | 1.21 | 1.55 | 1.28 | 1.11 | 1.47 |
| Longstanding illness | 1.95 | 1.66 | 2.28 | 1.73 | 1.46 | 2.05 |
| **Vulnerable to poverty 2** | | | | | | |
| Raised Depression | 2.27 | 1.84 | 2.80 | 1.85 | 1.46 | 2.35 |
| Overweight | 1.45 | 1.30 | 1.62 | 1.19 | 1.04 | 1.36 |
| Longstanding illness | 1.53 | 1.32 | 1.78 | 1.38 | 1.16 | 1.64 |
| **Persistent poverty** | | | | | | |
| Raised Depression | 5.52 | 4.48 | 6.79 | 3.90 | 2.98 | 5.12 |
| Overweight | 2.23 | 1.93 | 2.58 | 1.61 | 1.33 | 1.95 |
| Longstanding illness | 2.94 | 2.50 | 3.44 | 2.13 | 1.73 | 2.60 |

**Abbreviations:** OR odds ratio, LCI lower confidence interval (95), UCI upper confidence interval (95), SDQ strengths and difficulties questionnaire.
**Adjusted** by mother age, education (degree or higher qualification or no degree), health status at 9 months (chronic illness, BMI), depression status at 9 months (raised depression score), home-owner, household social class and composition. Sensitivity analysis using multiple imputation, (20 draws). See bottom of Table 1 for (n) of each poverty group and outcome.

**Table S13:** Sensitivity analysis – reporting incidence of risk ratio in the association of predicted poverty trajectories from 9 months to 9 years and child outcomes at 9 years using the Irish Growing Up in Ireland – Infant Cohort

| | | Never in Poverty | Vulnerable to Poverty 1 | | | Vulnerable to Poverty 2 | | | Persistent Poverty | | |
|---|---|---|---|---|---|---|---|---|---|---|---|
| | | | **IRR** | **CI** | | **IRR** | **CI** | | **IRR** | **CI** | |
| **Total SDQ Difficulties – Parent** | **UA** | Ref | 2.23 | 1.65 | 3.02 | 2.33 | 1.78 | 3.05 | 3.84 | 2.93 | 5.05 |
| | **A** | | 1.91 | 1.42 | 2.57 | 1.49 | 1.09 | 2.03 | 1.93 | 1.38 | 2.70 |
| **Total SDQ Difficulties - Teacher** | **UA** | Ref | 1.27 | 1.03 | 1.56 | 1.76 | 1.47 | 2.10 | 2.69 | 2.25 | 3.21 |
| | **A** | | 1.22 | .99 | 1.50 | 1.41 | 1.15 | 1.73 | 1.75 | 1.40 | 2.20 |
| **BMI – Overweight** | **UA** | Ref | 1.22 | 1.06 | 1.42 | 1.49 | 1.31 | 1.70 | 1.89 | 1.64 | 2.18 |
| | **A** | | 1.14 | .99 | 1.32 | 1.24 | 1.07 | 1.44 | 1.47 | 1.24 | 1.73 |
| **Longstanding Health Condition** | **UA** | Ref | 1.25 | 1.10 | 1.43 | 1.13 | 1.00 | 1.29 | 1.45 | 1.26 | 1.67 |
| | **A** | | 1.23 | 1.08 | 1.40 | 1.13 | .98 | 1.30 | 1.36 | 1.15 | 1.61 |

**Abbreviations:** Incidence of Risk Ratio, CI Confidence Intervals, Ref reference group (never in poverty), SDQ Strengths and Difficulties Questionnaire, BMI Body Mass Index, UA – Unadjusted Model, A – Adjusted Model (mother age, education (degree or higher qualification or no degree), health status at 9 months (chronic illness, BMI), depression status at 9 months (raised depression score), home-owner, household social class and composition, and child birthweight (<2500g (i.e., low birth weight) or >2500g) and pregnancy gestation (<37 (i.e., preterm) or >37 weeks)). See bottom of Table 1 for (n) of each poverty group and outcome.

**Table S14:** Sensitivity analysis – reporting incidence of risk ratio in the association of predicted poverty trajectories mother outcomes after 9 years using the Irish Growing Up in Ireland – Infant Cohort.

| | | Never in Poverty | Vulnerable to Poverty 1 | | | Vulnerable to Poverty 2 | | | Persistent Poverty | | |
|---|---|---|---|---|---|---|---|---|---|---|---|
| | | | **IRR** | **CI** | | **IRR** | **CI** | | **IRR** | **CI** | |
| **Depression Raised Score** | **UA** | Ref | 2.14 | 1.66 | 2.75 | 2.30 | 1.80 | 2.94 | 4.65 | 3.71 | 5.82 |
| | **A** | | 1.92 | 1.49 | 2.49 | 1.78 | 1.36 | 2.33 | 3.01 | 2.26 | 4.01 |
| **BMI - Overweight** | **UA** | Ref | 1.13 | 1.05 | 1.21 | 1.18 | 1.10 | 1.26 | 1.36 | 1.26 | 1.46 |
| | **A** | | 1.09 | 1.02 | 1.17 | 1.08 | 1.00 | 1.16 | 1.14 | 1.05 | 1.25 |
| **Longstanding Health Condition** | **UA** | ref | 1.62 | 1.40 | 1.88 | 1.41 | 1.21 | 1.65 | 2.30 | 1.99 | 2.67 |
| | **A** | | 1.44 | 1.25 | 1.67 | 1.28 | 1.08 | 1.51 | 1.71 | 1.42 | 2.06 |

**Abbreviations:** OR Odds Ratio, CI Confidence Intervals, Ref Reference group (never in poverty), BMI Body Mass Index, UA – Unadjusted Model, A – Adjusted Model (mother age, education (degree or higher qualification or no degree), health status at 9 months (chronic illness, BMI), depression status at 9 months (raised depression score), home-owner, household social class and composition)). See bottom of Table 1 for (n) of each poverty group and outcome.

**Figures**

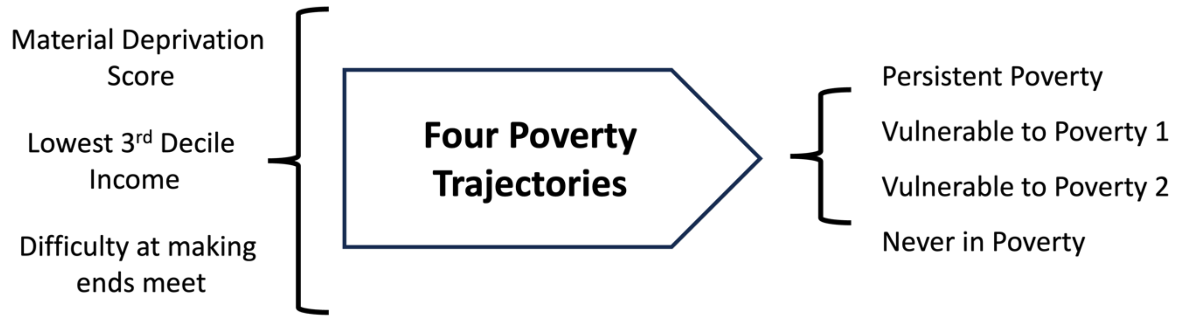


**Figure S1:** Group based trajectory of poverty using the Irish Growing Up in Ireland - Infant Cohort

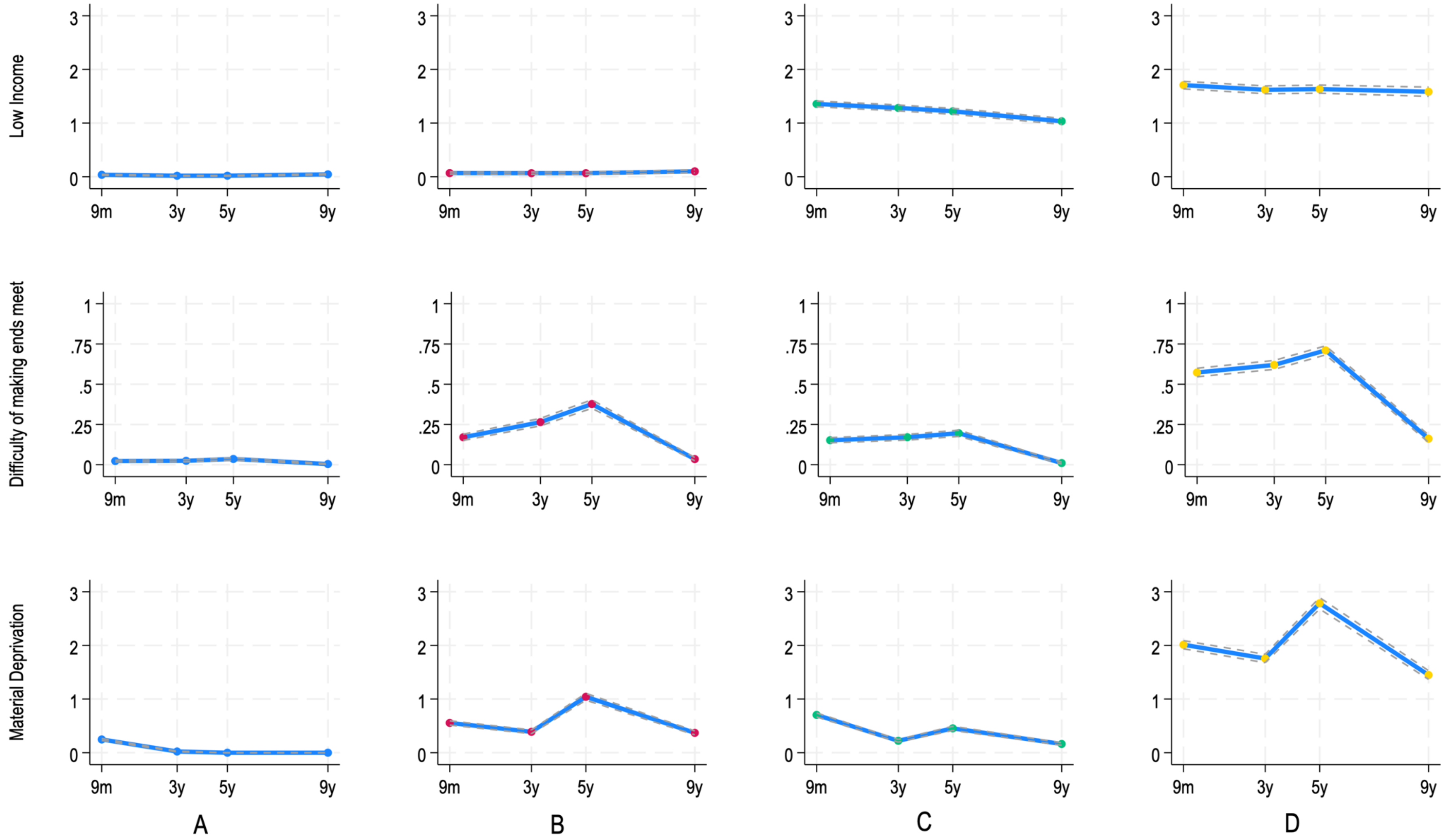


**Figure S2:** Group based trajectory of poverty of lowest third decile income, ease to meet bills (difficult or very difficult), and number of material deprivations (0-6) into four trajectories: (A) never poverty (43.1%), (B) vulnerable to poverty: material/perceived>monetary (16.1%), (C) vulnerable to poverty: monetary>material/perceived (25.6%) and (D) persistent poverty (15.2%) using the Irish Growing Up in Ireland - Infant Cohort.

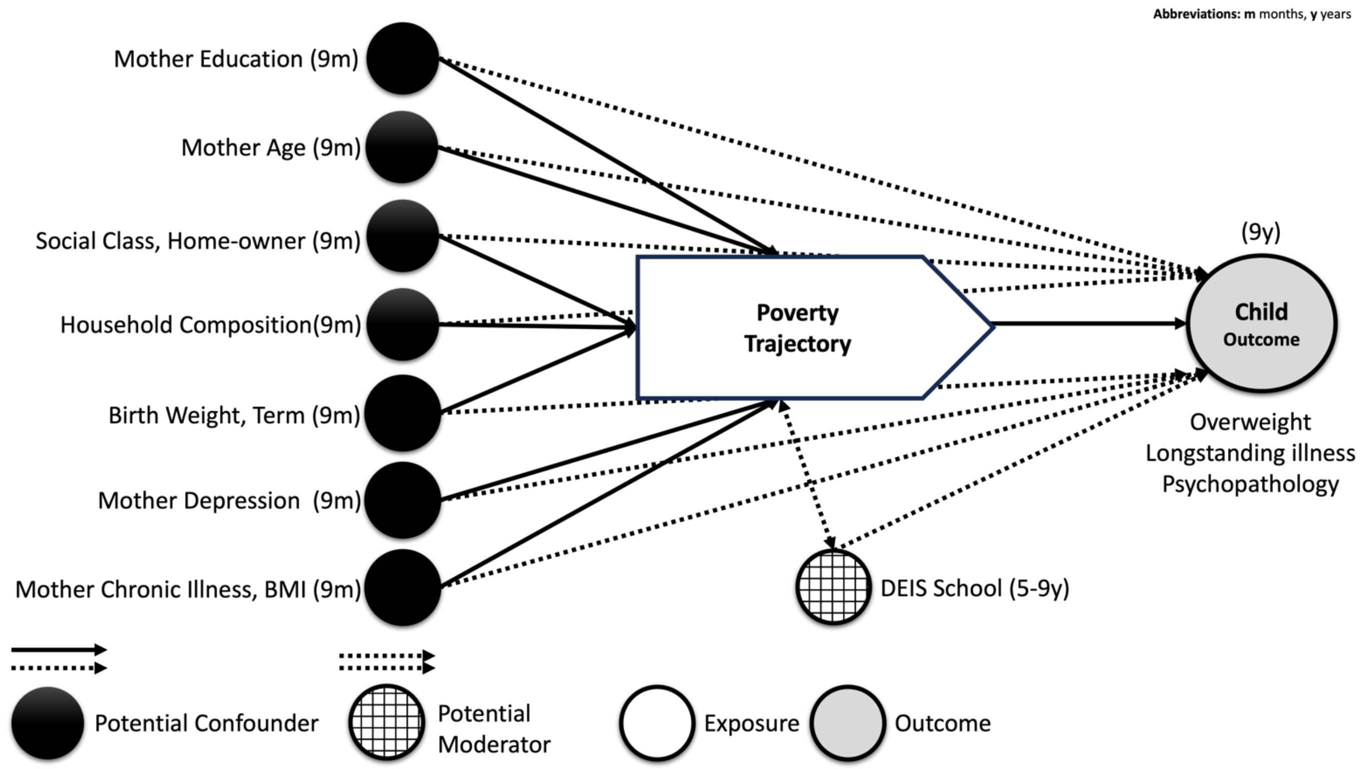


**Figure S3:** Directed Acyclic Graph (DAG) of association between poverty trajectory and child outcomes using the Irish Growing Up in Ireland - Infant Cohort

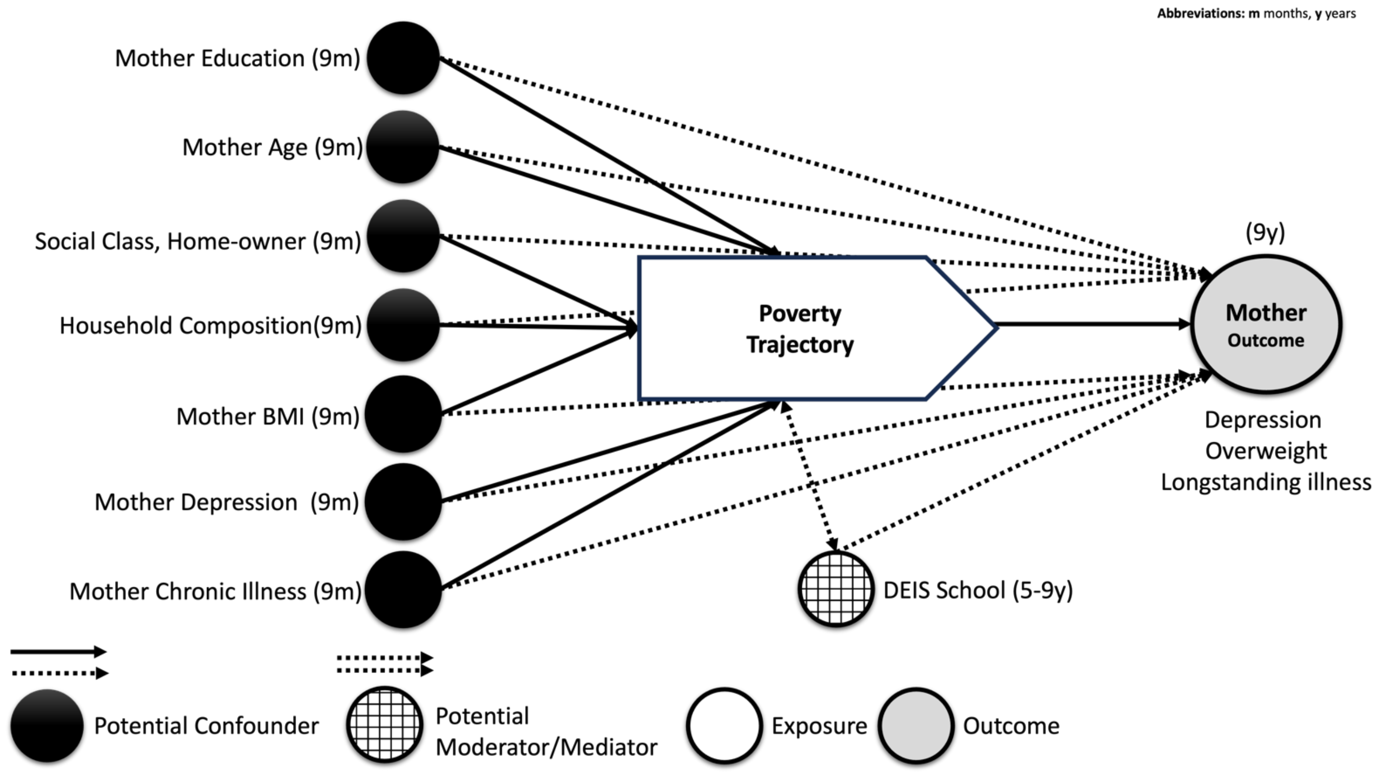


**Figure S4:** Directed Acyclic Graph (DAG) of association between poverty trajectory and mother outcomes using the Irish Growing Up in Ireland - Infant Cohort

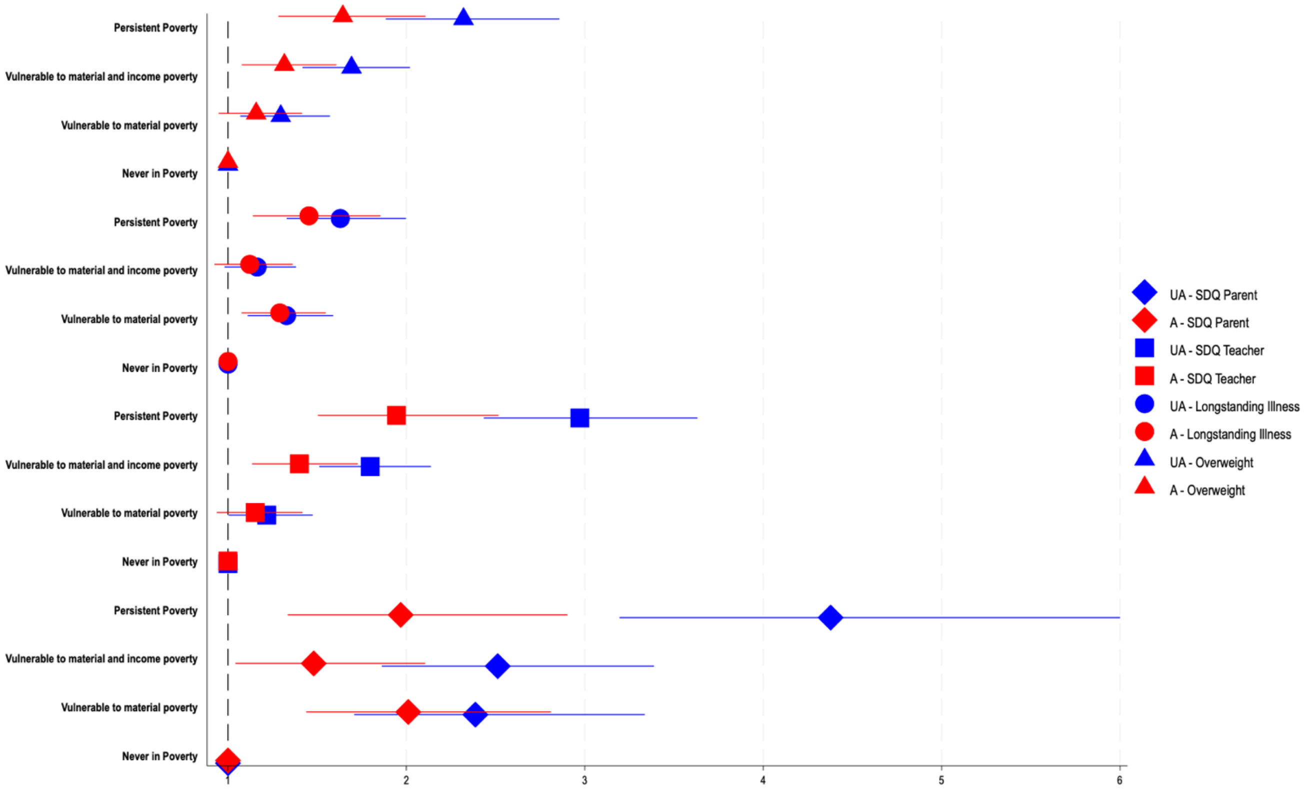


**Figure S5:** Associations of poverty trajectories (never in poverty, vulnerable to material poverty (group 1), vulnerable to material and income poverty (group 2) and persistent poverty) and child outcomes (total strengths and difficulties (SDQ) scores by parent and by teacher, longstanding illness, and overweight) at age 9 in the Irish Growing Up in Ireland Study – Infant cohort. Models were unadjusted (UA) and adjusted (A) for mother age, education (degree or higher qualification or no degree), health status at 9 months (chronic illness, BMI), depression status at 9 months (raised depression score), home-owner, household social class and composition, and child birthweight (<2500g (i.e., low birth weight) or >2500g) and pregnancy gestation (<37 (i.e., preterm) or >37 weeks).

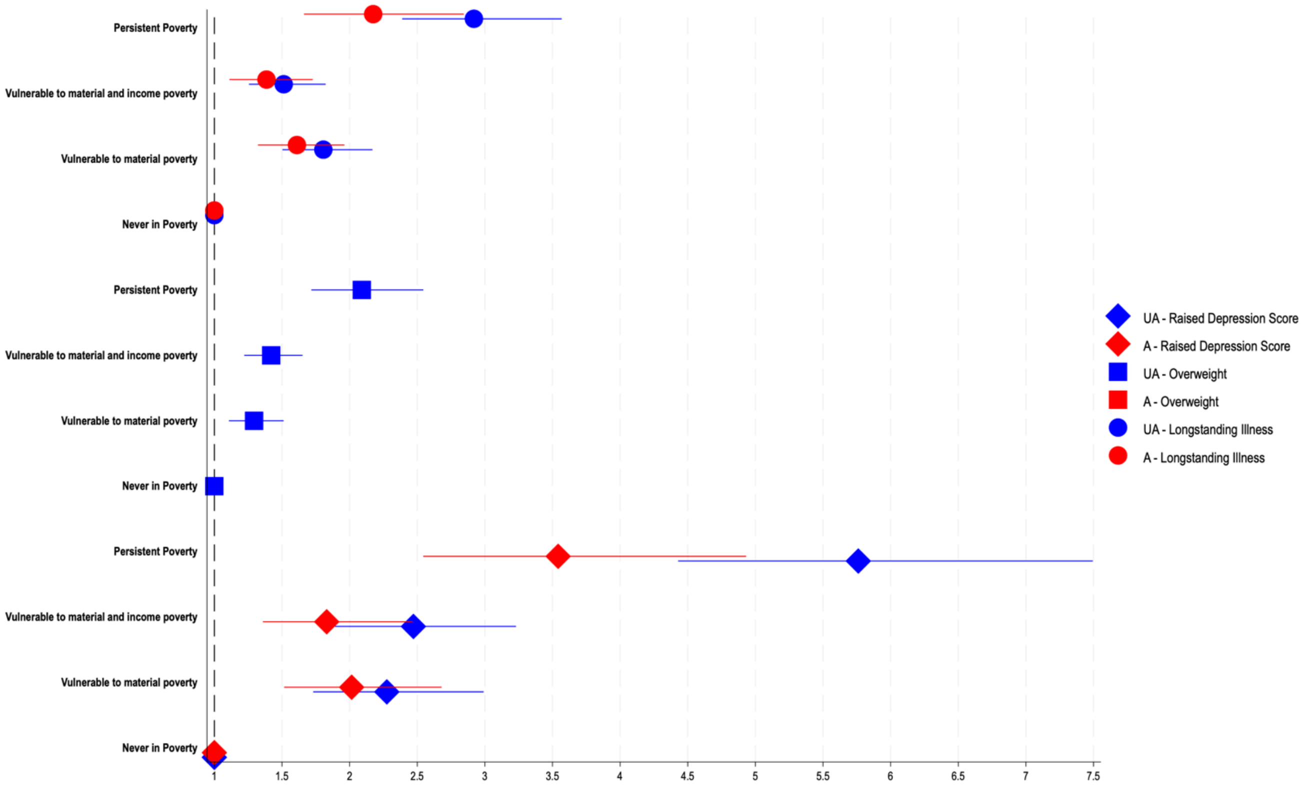


**Figure S6:** Associations of poverty trajectories (never in poverty, vulnerable to material poverty (group 1), vulnerable to material and income poverty (group 2) and persistent poverty) and mother outcomes (raised depression score, longstanding illness, and overweight) at age 9 in the Irish Growing Up in Ireland Study – Infant cohort. Models were unadjusted (UA) and adjusted (A) for PCG age, education (degree or higher qualification or no degree), health status at 9 months (chronic illness, BMI), depression status at 9 months (raised depression score), home-owner, household social class and composition.
